# Functional MRS uncovers age-related alterations in cerebral lactate dynamics during emotional-cognitive engagement, revealing metabolic vulnerability in the dACC

**DOI:** 10.64898/2026.02.05.26345665

**Authors:** Edward Caddye, Joel Patchitt, Anouk Schrantee, William T Clarke, Itamar Ronen, Alessandro Colasanti

## Abstract

**Introduction:** Lactate plays dual roles in neuronal energy metabolism and signalling. The dorsal anterior cingulate cortex (dACC), a region with high baseline glycolytic activity implicated in psychiatric disorders, may exhibit dynamic lactate responses to graded cognitive-emotional demands. Because mitochondrial function declines with age, aging may model whether fMRS-derived lactate dynamics can detect latent neurometabolic vulnerabilities.

**Methods:** Using fMRS, we monitored dACC metabolite changes in 34 healthy participants (aged 21–69) during an emotional face-processing task with escalating cognitive-emotional workload. The paradigm comprised a 2-minute baseline, 10-minute task of increasing intensity, and 10-minute recovery.

**Results:** dACC lactate increased significantly, tracking task intensity and peaking 19.5% above baseline at maximum cognitive load (z = 2.66, p = 0.004). The response showed both linear task-related increases (z = 2.08, p = 0.02) and a quadratic inverted-U profile (z = 2.72, p = 0.004). Total creatine, total NAA and Glx (Glutamate+Glutamine) exhibited no task-dependent changes. Age influenced task-period lactate AUC (z = 2.19, p = 0.014). Participants over 40 exhibited greater peak responses (54% vs 28%), steeper upslopes (14% vs 7% per block), and larger AUC (155% vs 16%) than those under 40. Sex differences were also observed. Baseline lactate did not correlate with age.

**Conclusions:** dACC lactate dynamics are sensitive to cognitive-emotional demand, with evidence of age-and sex-dependent modulation. The dissociation between static and dynamic measures establishes a metabolic stress-testing paradigm for detecting latent neuroenergetic vulnerabilities, supporting fMRS utility for probing mitochondrial function in health and psychiatric disorders.

## Introduction

Lactate, once misclassified as a waste product of anaerobic metabolism, plays dual roles in neuronal energy metabolism and signalling (1). Its regulation depends on continuous production and clearance throughout the brain and body (2). In the brain, glucose is primarily metabolised via glycolysis during neuronal activation. The rationale for this upregulation under aerobic conditions is not fully understood: earlier theories suggested that glycolysis had a temporal advantage over oxidative phosphorylation (3), whilst a more recent theoretical model posed that glycolysis during aerobic conditions may support neuronal signalling in thin axons that are highly information-efficient but lack mitochondria (4). Lactate is subsequently released into circulation and cleared by increased cerebral blood flow and the glymphatic system. The astrocyte-neuron lactate shuttle (ANLS) describes how astrocytes use aerobic glycolysis to support neurons metabolically (5). While this highlights a supportive role during activation, the function of lactate as a primary neuronal fuel in the adult brain remains debated (6). Lactate dynamics are modulated by cortical arousal (7), catecholamines (8,9), and are involved in long-term memory formation (10), suggesting that changes in lactate reflect, and may influence, bioenergetic and neuromodulatory states relevant to stress and psychiatric pathophysiology.

Lactate dynamics have long been studied in exercise physiology (11), but their relevance to psychiatry is now emerging. Early findings showed altered peripheral lactate responses in anxiety (12), and recent evidence supports lactate as a biomarker of mitochondrial dysfunction in mood disorders (13). Elevated brain lactate is a shared endophenotype across several psychiatric conditions (14), and there is growing interest in its role as a potential therapeutic target and biomarker in the growing field of metabolic psychiatry (15–17).

Functional magnetic resonance spectroscopy (fMRS) offers a non-invasive, time-sensitive method to monitor brain metabolism, and is particularly relevant to psychiatric research. While the Blood Oxygenation Level Dependent (BOLD) contrast, which underpins fMRI can offer higher spatial and temporal resolution, its signal reflects a complex mix of vascular, metabolic, and neuronal activity that cannot easily be disentangled (18). Early MRS studies relied on static, resting-state lactate measures, with few exploring dynamic changes (19–22); most focused on glutamate or GABA responses to motor or visual stimuli, often overlooking lactate (23). Although preliminary findings suggest altered lactate dynamics in the visual cortex in panic disorder (24), or in response to exercise (25), recent work has demonstrated elevated dACC lactate at rest in chronic fatigue syndrome (26), and task-dependent lactate flux abnormalities in the precuneus in bipolar disorder (27), dynamic lactate responses to cognitive-emotional challenge in the anterior cingulate cortex (ACC) remain uncharacterised.

The ACC is an ideal brain region for studying lactate dynamics in the context of mood disorders due to its high baseline glycolytic activity and its central role in integrating emotion, cognition, and behaviour (28–30), and its pathophysiological relevance to bipolar disorder and depression (31–33), which have been linked to neurometabolic alterations of the ACC (34,35). Intracranial ACC stimulation elicits physiological arousal and motivational responses, consistent with its role in energy allocation and drive (36). Recent findings show ACC lactate levels correlate with effort-based decision-making, implicating lactate in the encoding of energy cost and motivational salience (37). Our study focuses on the dorsal ACC (dACC), a region involved in emotional appraisal, cognitive control, and error monitoring (38,39), particularly suitable for MRS acquisitions, and reliably engaged by emotionally salient fMRI paradigms in both patients and healthy controls (40). To our knowledge, no prior studies have examined lactate dynamics in the dACC during emotionally engaging tasks. This study addresses that gap using fMRS to monitor task-related metabolic changes under increasing cognitive-emotional load.

To assess whether lactate dynamics could serve as a proxy for mitochondrial function, we investigated whether lactate responses are sensitive to age-related changes in neurometabolism. Mitochondrial function declines with age (41), making aging a useful model for testing the sensitivity of fMRS-derived lactate measures. Recent work has suggested that brain lactate production at rest decreases by 7% per decade on average (42), but few, or perhaps none, have assessed how aging affects dynamic responses to task-based activation with fMRS. Characterising age-related shifts in lactate dynamics may provide early markers of mitochondrial inefficiency or dysfunction, a metabolic parameter relevant to both cognitive decline and psychiatric vulnerability.

The aims of this study were twofold: To investigate dynamic changes in dACC brain lactate during emotionally engaging cognitive effort, and to evaluate their sensitivity to the effects of aging in healthy participants. We hypothesised that: (1) lactate concentrations in the dACC would increase during the task with a physiological profile, reflecting increasing task intensity and metabolic demand, followed by recovery; and (2) older participants would exhibit greater peak lactate accumulation and slower recovery dynamics compared to younger participants, reflecting age-related reductions in mitochondrial efficiency.

## Methods and Materials

### Participants

Participants were scanned at the Clinical Imaging Sciences Centre (CISC) at the University of Sussex. Datasets were obtained from 45 healthy participants with normal or corrected-to-normal vision with no MRI contraindications. Participants with self-reported current or residual diagnoses of physical, neurological and/or psychiatric conditions were excluded. The study obtained ethical approval by the Research Governance and Ethical Committee of BSMS (ER/BSMS9YCR/1), and written and informed consent was obtained prior to participant involvement.

### MR acquisition

Imaging and spectroscopy data were obtained at 3 tesla (Magnetom Prisma, Siemens Healthineers, Germany). A T1-weighted (T1W) anatomical scan was acquired for each participant (TR = 2300 ms; TE = 2.19 ms; TI = 920 ms; Flip Angle = 9 degrees; FOV = 256 mm; voxel size = 1x1x1 mm^3^). This T1W image was used to place a volume of interest (VOI) mask measuring 30 x 20 x 15 mm^3^ in the dACC. On the coronal and axial midline, the bottom of the VOI was aligned anteriorly and superiorly to the corpus callosum, with the bottom corner aligned to the genu (Fig. 1A).

**Figure 1.**
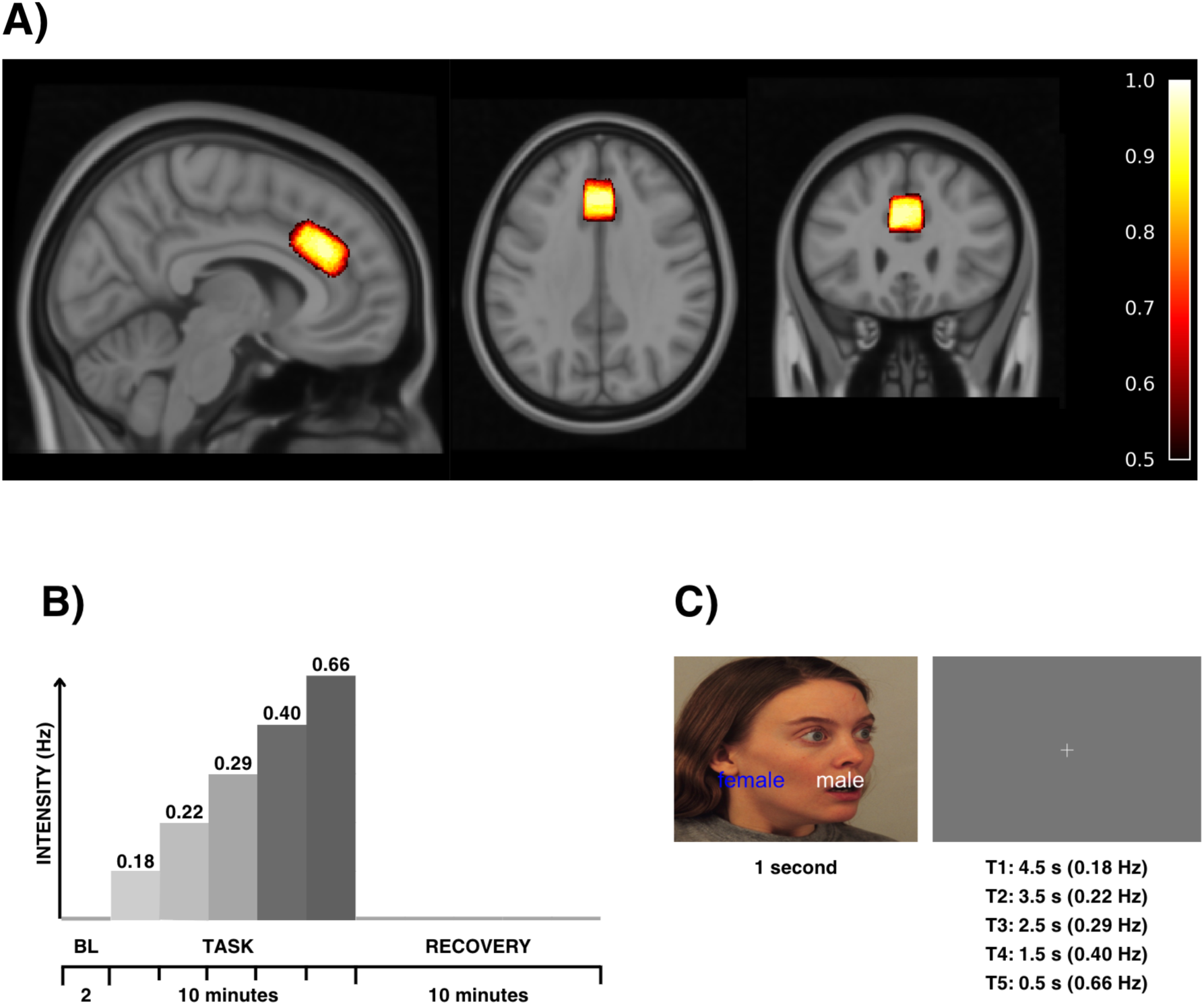
Functional magnetic resonance spectroscopy (fMRS) paradigm: **(a)** Spectroscopic data were acquired from the dorsal anterior cingulate cortex (dACC), from a 30 × 20 × 15 mm volume of interest. Sagittal, axial, and coronal views show the spatial distribution of individual spectroscopy voxel placements warped to standard MNI space. The colour scale reflects the proportion of participants with voxel coverage at each location, ranging from 50% to 100% overlap, on the MNI152 1 mm T1W brain template. **(b)** The MRS sequence spanned 22 minutes, during which time a task was presented to the participant comprising a 2-minute baseline (BL), a 10-minute task period, and a 10-minute recovery period. The task period was further divided into five 2-minute intervals. Each 2-minute block was designed to expose the participants to an incremental increase in intensity of workload (indicated by the darkness and height of the shaded blocks). **(c)** The task, coded for in PsychoPy, involved implicit emotional face processing and rapid decision-making. Emotional faces were presented for 1 second, and participants indicated within that time via button press in the right hand whether each face was male or female. Between face presentations, a fixation cross appeared, with the duration of this rest period progressively decreasing from 4.5 seconds to 0.5 seconds and the face presentation frequency (Task intensity) progressively increased from 0.18 to 0.66 Hz.

MRS data were acquired with a semi-LASER sequence ((43,44) Release 2016-12, CMRR Spectroscopy Package, University of Minnesota) using GOIA-WURST adiabatic inversion pulses, VAPOR water suppression and a 6-band outer volume suppression (OVS) (TR = 3000 ms; TE = 30 ms; bandwidth = 2.5 kHz; 2048 points, 448 dynamics) (43). Four unsuppressed water spectra were obtained for eddy current correction and water scaling, two at the beginning and two at the end of the data series. Calibration of the 90-degree excitation pulse and flip angle for the water suppression were performed automatically (44). B0 shimming was performed using a 4-stage FASTESTMAP routine (45).

### Experimental Design

This study utilised an fMRS design to investigate temporal changes in metabolites during an emotional-cognitive task, adapted from a paradigm previously shown to elicit altered hemodynamic responses in the dACC in bipolar disorder (40), simulating incremental increases in cognitive workload analogous to physical stress testing (11).

In the MRI scanner, stimuli were projected onto a screen behind the participants’ heads and viewed using a mirror attached to the head coil. During the initial T1W acquisition, participants were permitted to relax and close their eyes. Prior to MRS acquisition, participants were instructed to open their eyes and fixate on a central crosshair throughout the calibration sequences to ensure consistent visual engagement, and subsequently to keep their eyes open for the entirety of the MRS sequence, including task and baseline periods. After T1W acquisition and VOI mask placement (Fig. 1A), resulting in at least 10 minutes of rest inside the scanner, dACC spectroscopy data were acquired over a 22-minute period.

A total of 209 single-face colour images displaying task-irrelevant emotional expressions of varying intensity (46) were presented in random order during the task (Fig. 1B, C). Participants were instructed to discern the gender of each face using a right-hand button press. The index finger selected the left-side option, and the middle finger selected the right. To increase task difficulty and reduce habituation, the screen position of the gender labels ("male" or "female") was randomized on each trial. A response was considered correct only if it was made within the 1-second window within which time the face was presented. Task accuracy was calculated as the percentage of 209 trials in which a correct gender identification was made within this response window. These data were available for 29 participants due to technical recording failures in 5 participants with quality datasets.

### MRS pre-processing

fMRS data processing and dynamic fitting was carried out using FSL-MRS (version 2.1.19) (47). Pre-processing steps involved data conversion to NIfTI-MRS using spec2nii (version 0.7.3) (48), phase and frequency alignment of dynamics (49), and eddy-current correction using the unsuppressed water (50). For quality control, these pre-processed data were averaged for each participant to produce a single time-averaged spectrum for inspection. For dynamic fitting (51), the non-averaged pre-processed datasets were carried forward. A basis set containing 19 metabolite spectra (ascorbate (Asc), aspartate (Asp), creatine (Cr), gamma-amino butyric acid (GABA), glucose (Glc), glutamine (Gln), glutamate (Glu), glycine (Gly), glycerophosphocholine (GPC), glutathione (GSH), inositol (Ins), lactate (Lac), N-acetylaspartate (NAA), NAA-glutamate (NAAG), phosphocholine (PCho), phosphocreatine (PCr), phosphoethanolamine (PE), scyllo-inositol (sIns), taurine (Tau), and a macromolecular baseline (Mac)) was simulated using FSL-MRS (44), with an additional lipid moiety at 1.3 ppm (MM13) (52). This basis set was optimised by fitting it to the average of all subject data, allowing metabolites to shift in frequency relative to each other, the relative shifts then inputted back into the basis set. FSL-MRS applies a linear combination model to the data. Briefly, the spectral basis set is shifted, broadened, and scaled to match the free induction decay signal in the spectral domain. The fitting procedure used a nonlinear least squares optimisation algorithm and accounts for nuisance parameters, including zero-order and first-order phase corrections, and incorporates a second order polynomial baseline.

### fMRS processing and analysis

For the temporal modelling of metabolite level changes, FSL-MRS fits a dynamic signal model to all transients simultaneously (51), enhancing precision in detecting stimulus-induced changes compared to block averaging (53). A design matrix was produced with nilearn (54) to model the response of each metabolite in the context of a general linear model (GLM), consisting of 11 two-minute blocks (STIM00 to STIM10) and regressors for linear drift and constant (Fig. 2A). The parameters included in the dynamic model include the concentration of the metabolites in our basis set in addition to several others (Fig. 2B).

**Figure 2.**
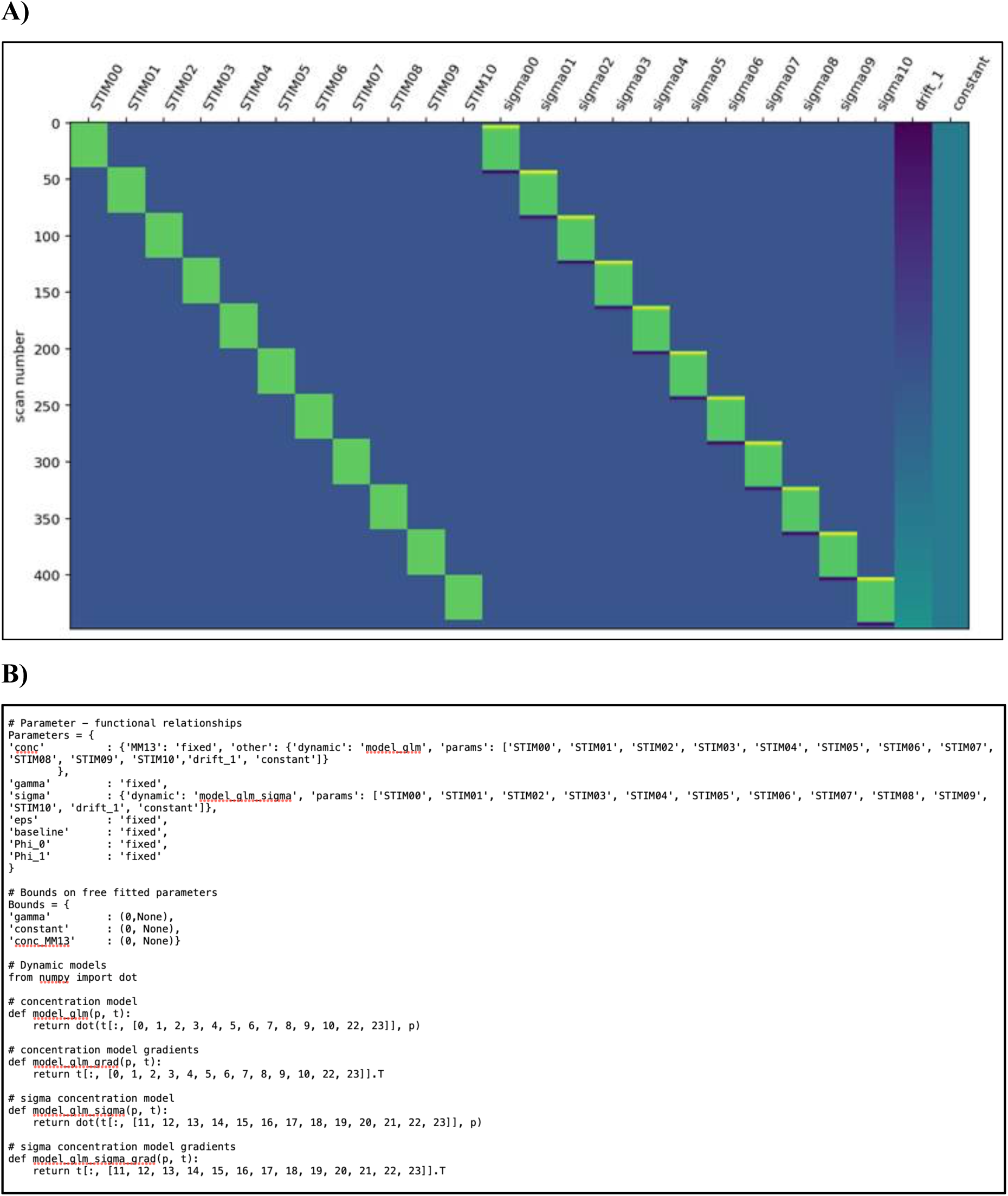
FSL-MRS pipeline dynamic fitting design matrix and configuration file: **(a)** Design matrix produced with nilearn. Each 2-minute block is defined in green and labelled across the X-axis from STIM00 to STIM10, and from sigma00 to sigma10 (convolved with the haemodynamic response function, HRF), alongside regressors that model linear drift and constant. The scan number from 0 to 448 dynamics is shown on the Y-axis. **(b)** Configuration file containing the parameters which are either fixed or dynamic. ‘Sigma’ (the Gaussian linewidth parameter) is modelled within the dynamic model and is convolved with the haemodynamic response function (HRF). The lipid MM13 is modelled as fixed, whereas other metabolites are modelled as dynamic.

We also chose to model ‘sigma’ (the Gaussian linewidth parameter) within the dynamic model, and convolve it with the haemodynamic response function (HRF), aligning with previous work (55,56), because changes in the BOLD signal over time can cause line narrowing in MR spectra due to alterations in T2* relaxation times (57,58). To optimise the dynamic fitting of lactate we included and modelled the lipid moiety MM13 as a fixed concentration parameter (Fig. 2B).

### Statistical Analysis

After dynamic fitting, beta values for each design matrix regressor (i.e., each STIM block, linear drift, and constant) were extracted for each metabolite and participant.

Our *a priori* primary objective was to characterise lactate dynamics in relation to task intensity. We defined three first-level temporal contrasts representing distinct, pre-specified aspects of the lactate response: (1) baseline-to-peak change (STIM00 vs. STIM05), (2) linear increase during the task (STIM01 to STIM05), and (3), quadratic inverted-U pattern across the full sequence (STIM01 to 10). To investigate the impact of age, we additionally sought to express the dynamic response using area under the curve (AUC) metrics: (4) baseline-corrected trapezoid AUC (btAUC) for the task (STIM01 to STIM05), (5) btAUC for recovery (STIM05 to STIM10) and (6), total btAUC (STIM01 to STIM10).

Statistical analyses were conducted using FSL-MRS (version 2.1.19). First-level contrast of parameter estimates (COPEs) were entered into the higher-level analyses using the fmrs_stats module, which implements mixed-effects modelling via FSL’s FLAME 1 algorithm. The higher-level design matrix included continuous age (demeaned), and sex. For each temporal contrast, we tested the following higher-level effects: (1) Continuous Age (demeaned); (2) Sex effect; (3) Sex x Age interaction (4) Group Mean. In a separate model we looked at whether baseline lactate influenced the dynamic trajectory, and as a final exploratory analysis, we added task accuracy as a 4^th^ variable (n = 29). Statistical significance for all contrasts was determined using Wald z-tests. Significance was inferred at p < 0.05.

For graphical visualisation, beta values were multiplied by the design matrix to generate subject-level time courses, with metabolite concentrations expressed as percentage change from baseline.

To better illustrate potentially biologically relevant differences in lactate trajectories that might depend on age, we created two groups (under and over the age of 40, young and old). The age grouping was defined based on prior evidence of metabolomic inflection points around the fourth decade of life (59). From individual time courses, we extracted several key response features for age-group comparisons: maximum % change, up slope, down slope, total AUC. Measures were % change, up slope (%/block) and AUC (% x block).

## Results

### Spectral Data Quality

A representative spectrum from a single participant averaged over the 448 transients is shown (Fig. 3A). Spectra from all 45 participants were inspected grossly, and as a result, 11 datasets were excluded from dynamic fitting and further analysis. These data were subject to lipid contamination that prevented accurate quantification of the lactate doublet (Fig. 2B).

**Figure 3.**
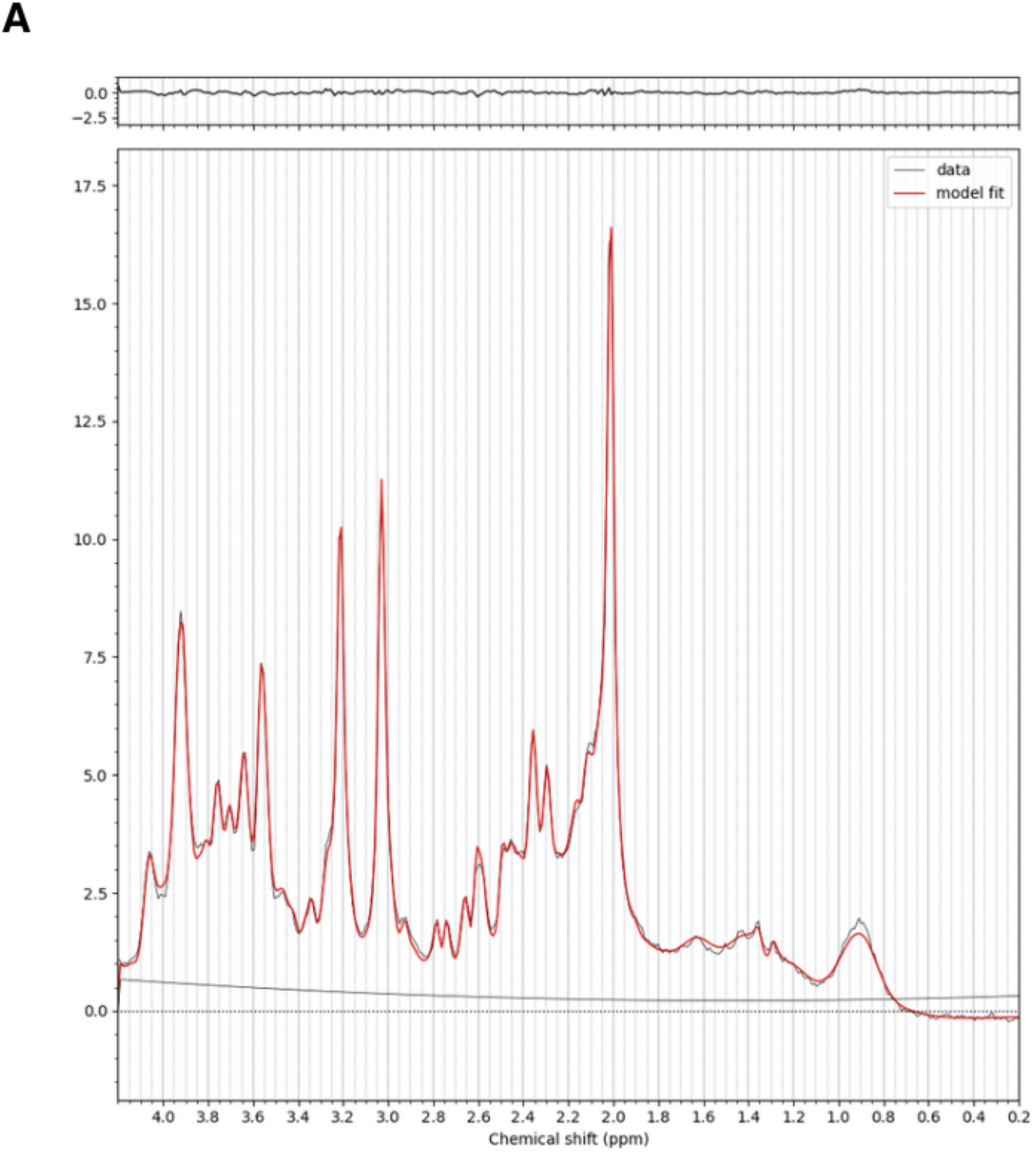
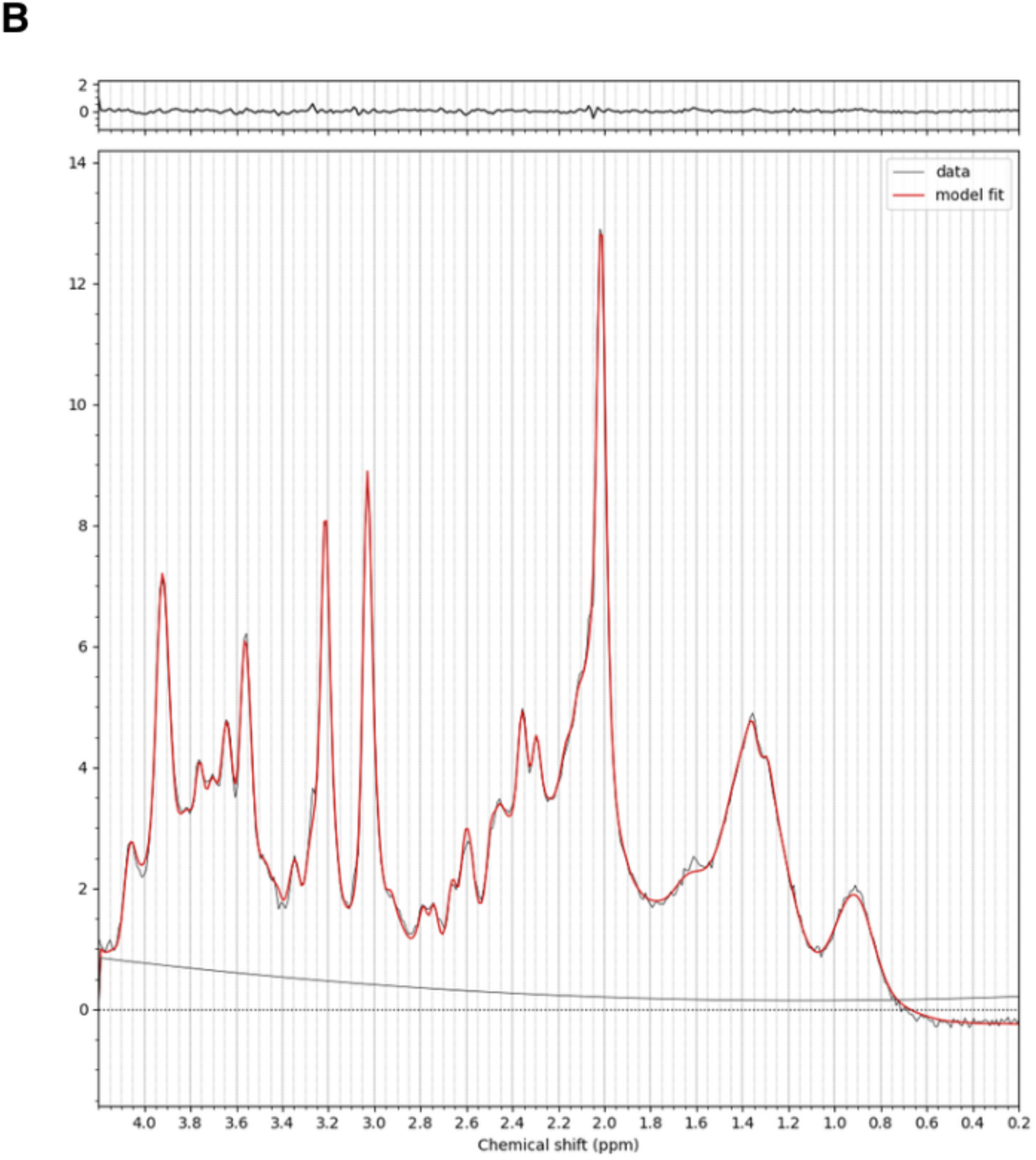
Average spectra from fitted FSL-MRS outputs from two participants: **(a)** Representative average spectrum from a single participant over 448 transients. **(b)** Example of an average spectra from one of the eleven excluded participant datasets with lipid contamination that prevents accurate quantification of the lactate doublet.

Hence, we included 34 participants (11 female, 23 male) ranging in age from 21 to 69 years (mean: 41.6 ± 17.0 years) in our analysis. Overall spectral quality was high: for NAA the Full Width at Half Maximum (FWHM) was 4.35 ± 0.54 Hz, and Signal-to-Noise Ratio (SNR) was 262 ± 99. Metabolite concentrations from these 34 participants were averaged over the total 448 transients (Average), along with the average across the 40 transients of the first block (Baseline) to ascertain the mean concentration (mM) and Percentage Cramer-Rao Lower Bound (%CRLB) for our metabolites of interest (Table 1).

**Table 1.** Quality control data. Mean ± SD concentration (mM) and %CRLB averaged across the first block of 40 transients (baseline) and the total 448 transients (average), reflecting the later temporal model regressors STIM00 and constant, respectively). Metabolites include lactate (Lac), glutamine (Gln), glutamate (Glu), aspartate (Asp), N-acetyl aspartate (NAA) and N-acetyl aspartylglutamate (NAAG).

| Block | Baseline | Average | Baseline | Average |
| --- | --- | --- | --- | --- |
| Metabolite | Mean ± SD (mM) | | %CRLB ± SD | |
| Lac | 1.02 ± 0.35 | 1.07 ± 0.29 | 26.6 ± 13.5 | 15.9 ± 4.8 |
| Gln | 3.03 ± 1.10 | 3.01 ± 0.66 | 11.9 ± 4.9 | 8.1 ± 2.2 |
| Glu | 13.1 ± 1.31 | 13.2 ± 1.23 | 3.0 ± 1.1 | 2.1 ± 0.6 |
| NAA | 13.6 ± 1.23 | 13.6 ± 1.19 | 1.7 ± 0.7 | 1.2 ± 0.4 |
| NAAG | 0.24 ± 0.17 | 0.24 ± 0.16 | 218 ± 361 | 158 ± 309 |

### Task-Dependent Neurometabolite Dynamics

Lactate levels in the dACC increased significantly during the task. A significant increase was observed from baseline (STIM00) to peak task intensity (STIM05) (z = 2.66, p = 0.004), with lactate levels rising on average by 19.5% at maximum task intensity (0.66 Hz) (Fig. 4). This task-dependent change was not dependent on baseline lactate concentration (z = -0.78, p = 0.78). Lactate exhibited a significant linear increase during the task execution (z = 2.08, p = 0.02) and an overall quadratic inverted-U profile (z = 2.72, p = 0.003), with lactate rising during task phases, peaking at maximum intensity, and returning toward baseline during recovery (Fig. 4). This was not the case for tCreatine (Phosphocreatine+Creatine), the glutamatergic pool (Glx, Glutamate+Glutamine) and tNAA (NAA+NAAG) which did not exhibit any significant task-dependent changes (Table 2, Fig. 4).

**Figure 4.**
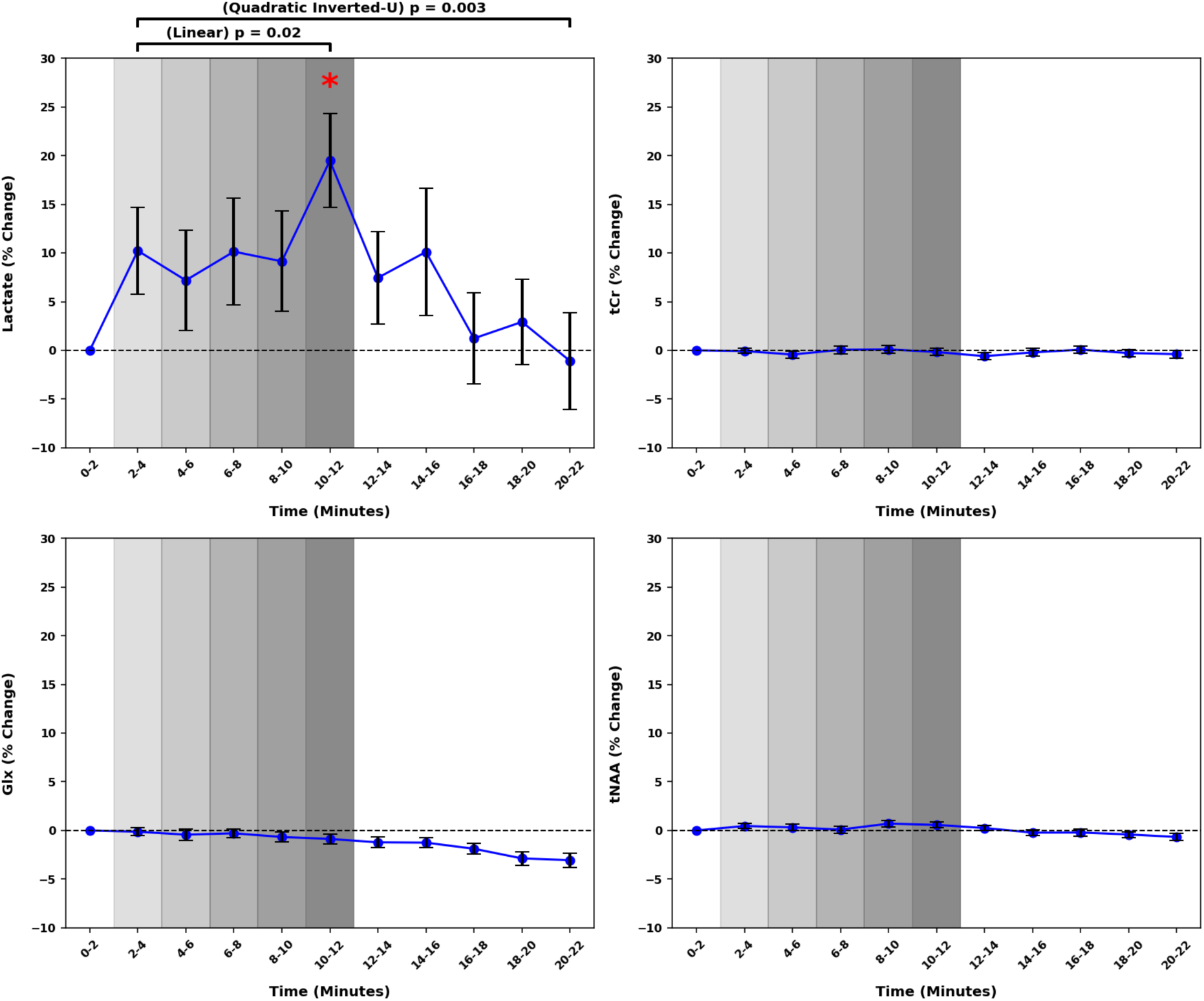
fMRS results: Average task-induced changes on metabolite levels in the dACC. Mean ± SEM as a percentage change from the first block (0-2 min) for 34 participants for Lactate, Total Creatine (PCr+Cr), Glutamatergic Pool (Glx, Glu+Gln) and Total N-Acetylaspartate Pool (tNAA, NAA+NAAG). Lactate exhibited a significant linear increase during the task period (STIM01 to 05; z = 2.08, p = 0.02), and the dynamics showed a significant quadratic (inverted-U) temporal profile (STIM01 to 10; z = 2.72, p = 0.003). A significant increase was observed from baseline (STIM00) to peak task intensity (STIM05) (z = 2.66, p = 0.004) for lactate and is indicated by red asterisk.

**Table 2.**
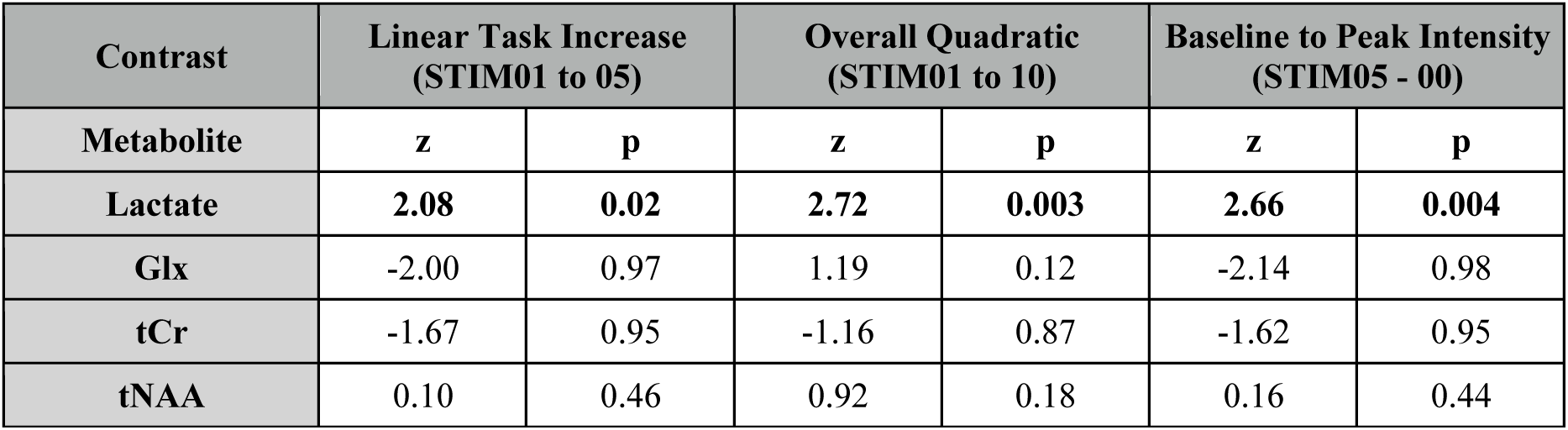
Statistical analysis of metabolite dynamics during an emotional-cognitive task. For lactate (a priori hypotheses), p-values are uncorrected. For other metabolites p-values are FDR-corrected across three metabolites per contrast. Wald z-statistics with p values are shown for Glx = Glutamate + Glutamine; tCr = Total Creatine (Creatine + Phosphocreatine); tNAA = total N-Acetylaspartate (NAA + NAAG).

| Contrast | Linear Task Increase (STIM01 to 05) |  | Overall Quadratic (STIM01 to 10) |  | Baseline to Peak Intensity (STIM05 - 00) |  |
| --- | --- | --- | --- | --- | --- | --- |
|  | z | p | z | p | z | p |
| Lactate | 2.08 | 0.02 | 2.72 | 0.003 | 2.66 | 0.004 |
| Glx | -2.00 | 0.97 | 1.19 | 0.12 | -2.14 | 0.98 |
| tCr | -1.67 | 0.95 | -1.16 | 0.87 | -1.62 | 0.95 |
| tNAA | 0.10 | 0.46 | 0.92 | 0.18 | 0.16 | 0.44 |

### Effect of Age on Lactate Dynamics

We observed no correlation between age and baseline lactate concentration (Pearson coefficient, r = 0.148, p = 0.41). There was also no correlation between age and grey matter tissue fraction (GM%) (r = -0.012, p = 0.948). In our group-level analysis, we investigated area-under-curve (AUC) differences for the task period (STIM01 to STIM05), recovery period (STIM05 to STIM10) and the total period (STIM01 to STIM10), as well as differences in the trajectory (STIM01 to STIM05) and the shape of the curve (quadratic inverted-U STIM01 to STIM10) for lactate dynamics. Strikingly, the effect of continuous age for the AUC task period was significant (z = 2.19, p = 0.014) as well as the total AUC (z = 1.86, p = 0.031) (Table 3). There was a significant effect of Sex on lactate dynamics, as well as an Age x Sex interaction, which was strongest for the linear response to the task (z = 2.58, p = 0.005) (Table 3).

**Table 3.** Effect of age (and biological sex) on lactate dynamics. Wald z tests and p values for each of the contrasts are shown for Age (continuous, demeaned), Sex and Sex x Age interaction. Baseline-corrected trapezoid area-under-curve (AUC) contrasts for the task (STIM01 to STIM05), recovery (STIM05 to STIM10) and total (STIM01 to STIM10) are shown. Other contrasts are for linear response to the task (STIM01 to STIM05), quadratic inverted-U response (STIM01 to STIM10), and the difference between baseline to peak intensity (STIM05 – STIM00). Statistical significance (p < 0.05) is highlighted in bold.

| Contrast | Age |  | Sex |  | Age x Sex |  |
| --- | --- | --- | --- | --- | --- | --- |
| Statistics | z | p | z | p | z | p |
| AUC Task | **2.19** | **0.014** | -0.38 | 0.65 | -2.90 | 1.0 |
| AUC Recovery | 0.83 | 0.20 | 0.23 | 0.41 | -1.75 | 0.96 |
| AUC Total | **1.86** | **0.031** | -0.32 | 0.63 | -2.76 | 0.99 |
| Linear Task | -1.93 | 0.97 | **1.88** | **0.03** | **2.58** | **0.005** |
| Quadratic Total | 0.36 | 0.68 | **2.73** | **0.003** | 0.21 | 0.41 |
| Baseline – Peak | -1.53 | 0.94 | **2.50** | **0.006** | **2.23** | **0.013** |

To characterise biologically relevant differences in specific aspects of lactate dynamics that might depend on age, we created two groups (under and over the age of 40, young and old). There was significant heterogeneity in the lactate response over time, but at the group level, there was a qualitative difference in the lactate response plotting relative change from the baseline block (Fig. 5).

**Figure 5.**
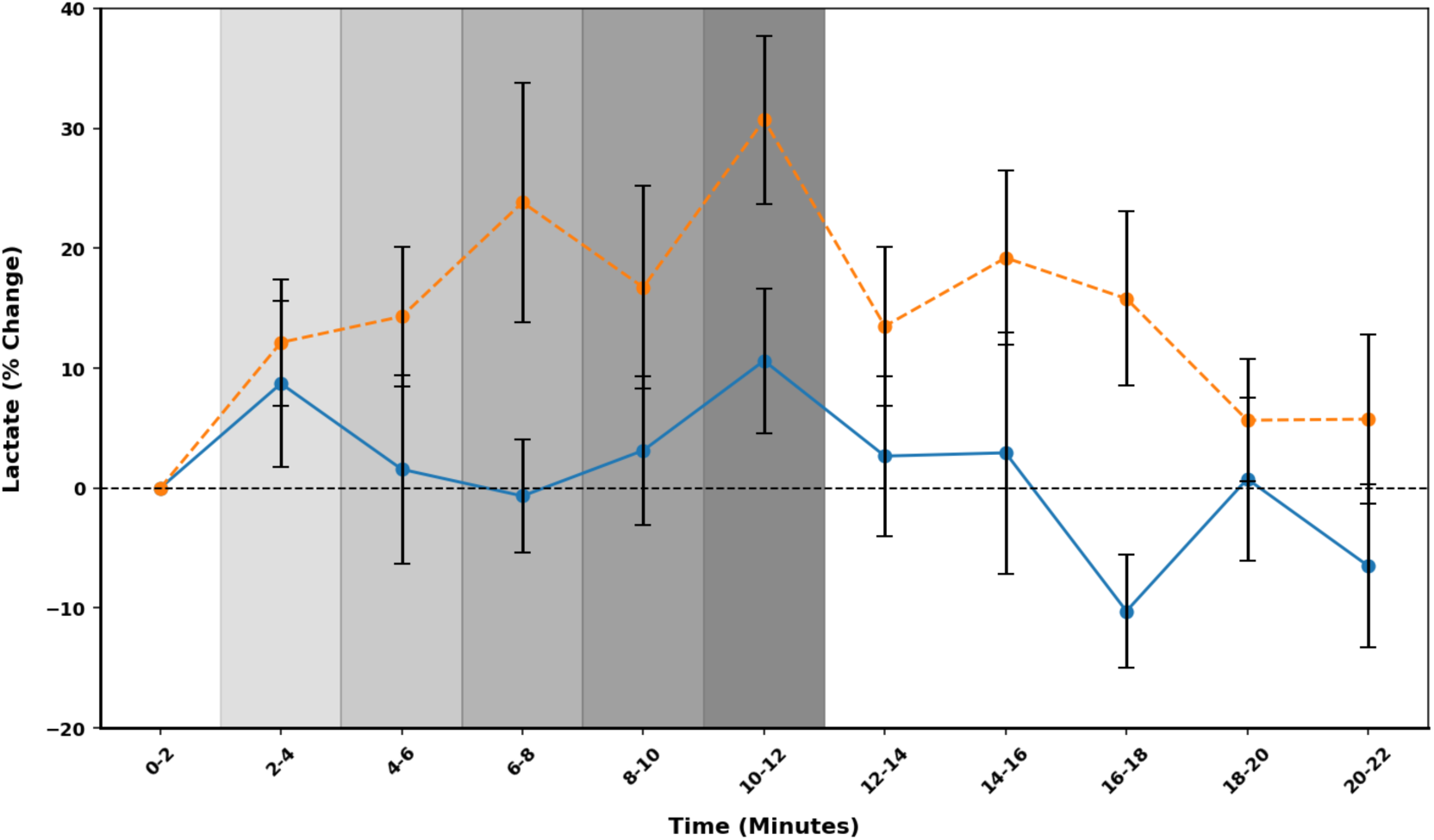
Lactate dynamics in participants under and over the age of 40 in response to an emotion-cognitive task in the dACC. Mean lactate ± SEM as a percentage change from the first block (0-2 min) in those over 40 (orange dashed, n = 15), and under 40 (solid blue, n = 19). The blocks corresponding with the task are shaded, which are progressively darker to highlight the reduction in time between face presentations and increase in intensity.

Key features of lactate dynamics were compared between groups (maximum % change, up slope, down slope, total AUC). The maximum % change, up slope (%/block) and AUC (% x block) differed significantly between young and old participants, which are summarised in Table 4.

**Table 4.**
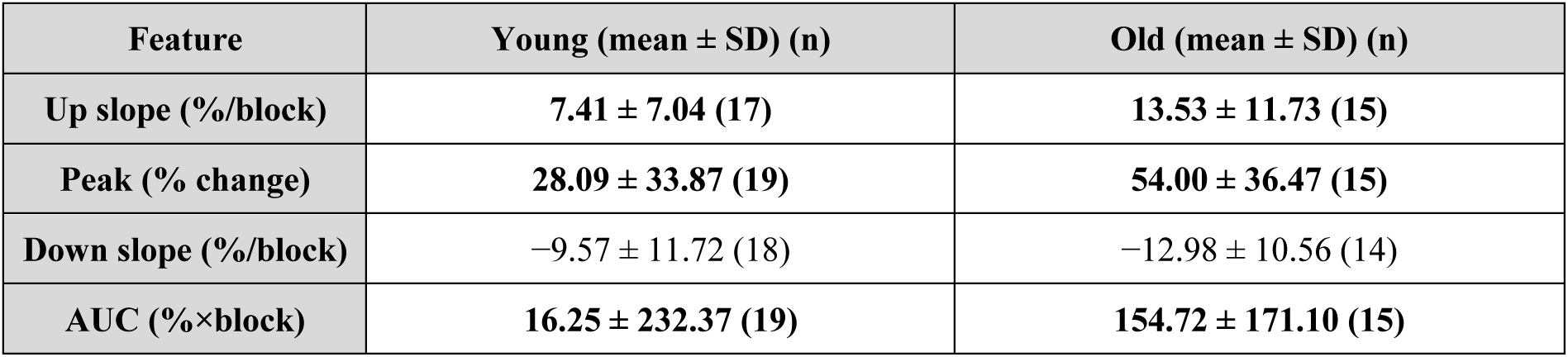
Lactate curve feature comparison between age groups (under and over 40). Peak amplitude and AUC were available for all participants. Up-slope and down-slope metrics were computed only for participants with valid peak positions and complete surrounding block values; therefore 2 younger participants were excluded from the up-slope calculation and 1 younger and 1 older participant were excluded from the down-slope calculation.

| Feature | Young (mean ± SD) (n) | Old (mean ± SD) (n) |
| --- | --- | --- |
| Up slope (%/block) | 7.41 ± 7.04 (17) | 13.53 ± 11.73 (15) |
| Peak (% change) | 28.09 ± 33.87 (19) | 54.00 ± 36.47 (15) |
| Down slope (%/block) | -9.57 ± 11.72 (18) | -12.98 ± 10.56 (14) |
| AUC (%×block) | 16.25 ± 232.37 (19) | 154.72 ± 171.10 (15) |

Finally, we identified that task accuracy, defined as the number of correct answers given within 1 second of the face presentation out of 209 trials, displayed a negative correlation with age (r = -0.541, p = 0.002). As an exploratory analysis, we re-ran our statistical analysis and added task accuracy as a regressor. In that limited dataset (n = 29), we found that task accuracy predicted the quadratic inverse-U response contrast for lactate (z = 2.11, p = 0.017).

## Discussion

Using fMRS, we demonstrate that dACC lactate responds dynamically to cognitive-emotional challenge, peaking 19.5% above baseline at maximum cognitive load. This response was not predicted by baseline lactate concentration at the individual level. Critically, baseline lactate showed no age or sex differences and did not correlate with age, yet metabolic challenge revealed significant age-dependent effects and sex-related modulation. This dissociation between static and dynamic measures establishes a neurometabolic stress-testing paradigm analogous to systemic exercise testing, wherein functional differences emerge only under metabolic demand.

The magnitude of the task-induced lactate response in the dACC is remarkably consistent with recently published work in an fMRS study in the motor cortex at 3 tesla (60). Here, the task-induced lactate increase exhibited both linear escalation during progressive task difficulty and a quadratic inverted-U profile, peaking at maximum intensity and returning toward baseline during recovery. This temporal pattern aligns with neuroenergetic models wherein task-evoked neural activity is primarily supported by upregulated glycolysis, with lactate serving as both metabolic substrate and signalling molecule (5). The tight coupling between lactate elevation and task intensity mirrors exercise-induced peripheral lactate responses (11), suggesting conserved principles of metabolic stress responses. Our observations indicate that fMRS-based evaluation of lactate dynamics offers a direct index of glycolytic flux during neural stimulation. Unlike other monitored metabolites (Glx, tCr, tNAA), which showed no task-dependent changes in this case, the unique sensitivity of lactate change to cognitive-emotional workload positions it as a candidate biomarker for neuroenergetic capacity and mitochondrial function.

Lactate dynamics can be decomposed into upslope, peak, downslope, and AUC, each reflecting distinct neurometabolic phenomena. Peak represents the net balance of lactate production via glycolysis and clearance at any given moment. In non-exercising healthy participants under normoxic conditions, observed lactate rises likely reflect *in situ* production rather than extra-cerebral import (61). AUC integrates both amplitude and duration of lactate elevation, reflecting overall metabolic engagement. Upslope likely reflects production rate, influenced by task characteristics, stress levels, and baseline metabolic state. Downslope may reflect metabolic consumption, export, or movement artefact. Unfortunately, single voxel fMRS cannot distinguish lactate changes between cellular or metabolic pools.

Biological age significantly influenced lactate response magnitude, particularly task AUC (z = 2.19, p = 0.014) and overall AUC (z = 1.86, p = 0.031). Participants over 40 exhibited greater peak increases (54% vs 28%), steeper upslopes (14% vs 7% per block), and larger AUC than those under 40, suggesting greater and faster glycolytic upregulation resulting in lactate accumulation. These findings align with established age-related declines in metabolic function (41,59), neurovascular (62) and neurometabolic coupling (63). Contrary to our *a priori* hypothesis, recovery dynamics did not differ significantly between age groups (p = 0.803), suggesting that lactate clearance mechanisms may be relatively preserved with age despite greater production. This dissociation between challenge response and recovery kinetics warrants further investigation with extended recovery periods or repeated-challenge paradigms.

Exaggerated lactate responses during cognitive challenge may reflect underlying mitochondrial inefficiency contributing to mental fatigue and reduced motivation. Historically, lactate infusions provoke panic in anxiety-prone individuals (64), though paradoxically may have anti-depressant effects (65). Recent work linking ACC lactate to effort-based decision-making suggests lactate serves dual roles as metabolic substrate and motivational signal (37). Interestingly, lactate in the ACC responds in a dose-dependent manner to ketamine infusion (56). The dissociation between metabolic demand (elevated lactate) and task performance in aging suggests that excessive lactate may index compensatory effort rather than efficient processing, or alternatively, an adaptive response to increased task difficulty. Recently, patients with bipolar disorder showed elevated precuneus lactate during emotional processing compared to controls (27), highlighting the importance of investigating task-dependent changes in patient populations.

Significant sex-dependent differences in lactate dynamics emerged, with the Sex × Age interaction suggesting sexually dimorphic metabolic responses to cognitive-emotional challenge. The interaction pattern showed that both linear task-related increases (z = 2.58, p = 0.005) and baseline-to-peak differences (z = 2.23, p = 0.013) varied as a function of both age and sex. Additionally, independent sex effects were observed for linear task responses (z = 1.88, p = 0.03), quadratic profiles (z = 2.73, p = 0.003), and baseline-to-peak changes (z = 2.50, p = 0.006), indicating that sex modulates lactate dynamics beyond age-related effects. This pattern may have implications for understanding cognitive vulnerability in aging and may be particularly relevant for understanding sex differences in psychiatric risk (66).

The biological mechanisms underlying these sex differences may involve the effect of estrogen on mitochondrial function (67), sex-specific patterns of neuromodulation in the dACC (68), or differential stress reactivity (69). Critically, these metabolic differences were not apparent at baseline, reinforcing that functional challenge paradigms reveal sex-related vulnerabilities masked in resting-state assessments. However, our small study sample, absence of hormonal profiling, menstrual cycle assessment, or menopausal status characterisation preclude definitive mechanistic interpretation. Nevertheless, the convergence of exaggerated metabolic responses with epidemiological patterns of psychiatric risk in older women suggests that sex-dependent metabolic aging may contribute to vulnerability for mood and anxiety disorders.

These findings establish proof-of-principle for dynamic fMRS as a translational tool for conceptualising therapeutic approaches in metabolic psychiatry. Analogous to cardiac stress testing, which unmasks latent coronary insufficiency not apparent at rest, neurometabolic stress-testing via fMRS may detect bioenergetic nuances before they manifest clinically. This framework potentially resolves a paradox: chronically elevated resting lactate may reflect mitochondrial impairment (70), whereas acute challenge-induced responses may index preserved metabolic resilience. The magnitude and recovery kinetics of lactate responses could thus serve as functional biomarkers with potential utility in staging illness severity, predicting treatment response, or monitoring metabolic interventions such as the ketogenic diet (71). They may also help with predicting mental fitness. The dissociation between lactate and glutamate dynamics further suggests metabolic specificity: while glutamate alterations reflect neurotransmitter imbalances, lactate dynamics may uniquely index energetic capacity and mitochondrial health. Given growing interest in mitochondrial dysfunction as a mechanism in mood disorders, bipolar disorder, and post-viral syndromes (72), fMRS-based metabolic stress-testing warrants investigation as a stratification tool for clinical trials of metabolic therapeutics.

Several limitations warrant consideration. First, we obtained data from a single 30 × 15 × 20 mm³ voxel, precluding assessment of regional or cellular specificity. Second, poor signal-to-noise for individual lactate measurements and lipid contamination (necessitating 24% participant exclusion) currently limit clinical scalability, though improved acquisition methods are under development. Third, we did not control for time of day, dietary habits, or MRI familiarity, factors likely influencing perceived stress. Nor did we correlate our brain lactate dynamics with systemic measures of physiology. Fourth, the 2-minute baseline could be extended to improve data quality. Fifth, the single-challenge paradigm with 10-minute recovery cannot fully characterise recovery kinetics; repeated-challenge designs would better distinguish acute capacity from recovery efficiency. Sixth, task accuracy lacked reaction time data, and participants could revise responses within the 1-second window; more granular behavioural metrics would better characterise effort-performance relationships. Finally, we did not assess menstrual cycle phase, menopausal status, or hormone levels, precluding mechanistic interpretation of sex effects. Despite these limitations, our findings establish fMRS-based metabolic stress-testing as a promising translational tool for probing mitochondrial function in health and disease.

## Conclusion

We demonstrate that dACC lactate serves as a dynamic marker of cognitive-emotional demand, with responses significantly modulated by age. Critically, baseline lactate showed no age or sex differences, yet metabolic challenge revealed age-dependent effects: a dissociation between static and dynamic measures analogous to cardiac stress testing, reinforcing recent work in bipolar disorder (27). Unlike other metabolites, which showed no task-dependent changes, the sensitivity of lactate positions it as a candidate theranostic biomarker in metabolic psychiatry. These findings establish challenge-based fMRS as a translational tool for detecting latent energetic vulnerabilities, with potential for staging illness severity, predicting treatment response, and monitoring metabolic interventions in psychiatric disorders characterised by mitochondrial dysfunction.

## Data Availability

All data produced in the present study are available upon reasonable request to the authors

## Acknowledgements

The MRS package was developed by Gülin Öz and Dinesh Deelchand and provided by the University of Minnesota under a C2P agreement. WTC is funded by Wellcome (225924/Z/22/Z). This research was funded in whole, or in part, by the Wellcome Trust [Grant numbers 225924/Z/22/Z]. For the purpose of open access, the author has applied a CC BY public copyright licence to any Author Accepted Manuscript version arising from this submission. ChatGPT5 by OpenAI was used to assist with the preparation of python scripts used throughout the data analysis and presentation. Claude Opus 4.5 by Anthropic was used to optimise readability, spelling, punctuation and grammar within the final draft.

## Financial Disclosures

WTC receives royalties from licensing of FSL to non-academic, commercial parties. All other authors report no biomedical financial interests or potential conflicts of interest.

## Notes

### Author Declarations

Ethics committee of Brighton and Sussex Medical School gave ethical approval for this work

